# Perspectives on building sustainable newborn screening programs for sickle cell disease: Experiences from Tanzania

**DOI:** 10.1101/2020.10.23.20218081

**Authors:** Daima Bukini, Siana Nkya, Sheryl McCurdy, Columba Mbekenga, Karim Manji, Michael Parker, Julie Makani

## Abstract

Prevalence of Sickle Cell Disease is high in Africa, with significant public health effects to the affected countries. Many of the countries with the highest prevalence of the disease also have poor health care system, high burden of infectious diseases with many other competing healthcare priorities. Though, considerable efforts have been done to implement newborn screening for Sickle Cell Disease programs in Africa but still coverage is low. Tanzania has one of the highest birth prevalence of children with Sickle Cell Disease in Africa. Also, it is one of many other African countries to implement pilot projects for Newborn Screening for Sickle Cell Disease to assess feasibility. Several efforts have been made afterwards to continue providing the screening services as well as comprehensive care for Sickle Cell Disease. Using qualitative methods, we conducted In- Depth Interviews and Focus Group Discussions with policy makers, health care providers and families to provide an analysis of their experiences and perspectives on efforts to expand and sustain Newborn Screening for Sickle Cell Disease and related comprehensive care services in the country. Findings have demonstrated both the opportunities and challenges in the implementation and sustainability of the services in low resource settings. A key area of strengthening is full integration of the services in countries’ health care systems to facilitate coverage, accessibility and affordability of the services. However, efforts at the local level to sustain the programs are encouraging and can be used as a model in other programs implemented in low resources settings.

## 2.0 Introduction

Sustaining implementation of health programs in limited resource settings has many challenges [1–5]. An example is implementation of vaccination programs in Low and Middle Income Countries (LMICs), which started with support through Global Alliance for Vaccine and Immunization (GAVI), Gates Foundation and Global funds [2, 6]. Efforts to build capacity for the programs to be owned by the local government were done in parallel during the implementation period and partly succeeded to build sustainable programs. However, immunization programs coverage has now dropped in Middle Income Countries after withdrawal of support from GAVI [7]. On the other hand, Human Immunodeficiency Virus (HIV) programs succeeded to be integrated in LMICs and now they are widely used as a model to build sustainable programs in resource limited settings [5]. These examples highlight both the potential and challenges of integrating once well-funded programs into local healthcare systems through local funding in LMICs.

Prevalence of Sickle Cell Disease (SCD) is high in Africa, with significant public health effects to the affected countries [8]. Many of the countries with the highest prevalence of the disease also have poor health care system, high burden of infectious diseases with many other competing healthcare priorities [9]. NBS for SCD is proven to be one of the most cost-effective approach in reducing morbidity and mortality associated with the disease [10]. Several countries in Africa with the highest burden of the disease have been developing pilot programs to implement NBS for SCD services [11–18].Though, considerable efforts have been done to implement the programs but coverage in the countries is still low [17–21]. Most of the programs are implemented through research funding by academic Universities in fewer selected health care facilities [22, 23]. Data on feasibility and cost effectiveness of the programs already exist from studies and pilot programs implemented in different countries of Africa [11–18, 24]. The question that still remain is how we can sustain implementation of the programs in low resource settings. Tanzania being one of the countries having the highest birth prevalence of children with SCD also implemented pilot studies on NBS for SCD to assess feasibility of the programs in the country [8, 16, 25]. There have been several initiatives since then in Tanzania to integrate NBS and the comprehensive care services. In this paper, we aim to share efforts and experiences of NBS for SCD in Tanzania to build a sustainable program in a resource constrained environment hoping to highlight both the opportunities and challenges.

## 3.0 Methodology

### 3.1 Study Settings

This study was implemented in health facilities located in Dar es Salaam and Mwanza regions. In Dar es Salaam, the study was implemented in Temeke (TRRH), Amana (ARRH) and Mwananyamala (MRRH) regional referral hospitals. TRRH was involved in the NBS for SCD pilot program of 2015/2016 and continued with phase II which started in November 2017 [16]. ARRH and MRRH were included in this study because at that time there were planned efforts to expand NBS for SCD in the two hospitals. A study done between 2004 to 2009 shows that Dar es Salaam has one of the highest population prevalence (3.5/1000) of registered SCD cases [9]. Bugando Medical Centre (BMC) located in Mwanza region is serving as the zonal referral hospital for the lake and western zone of Tanzania where, the annual births of children with SCD is estimated to be one of the highest in the world [26]. A pilot NBS for SCD program implemented at BMC between August to September 2014, reported 1.4% of children with SCD out of 919 children screened and 19.7% children with trait [27]. Inclusion of BMC was motivated by both prevalence of SCD in the region served by the hospital as well as their experiences in implementing NBS for SCD [26, 27]. Figure 1 (A) and (B) below are maps of Dar es Salaam and lake zone regions showing geographical locations of health facilities included in this study.

**Figure 1.**
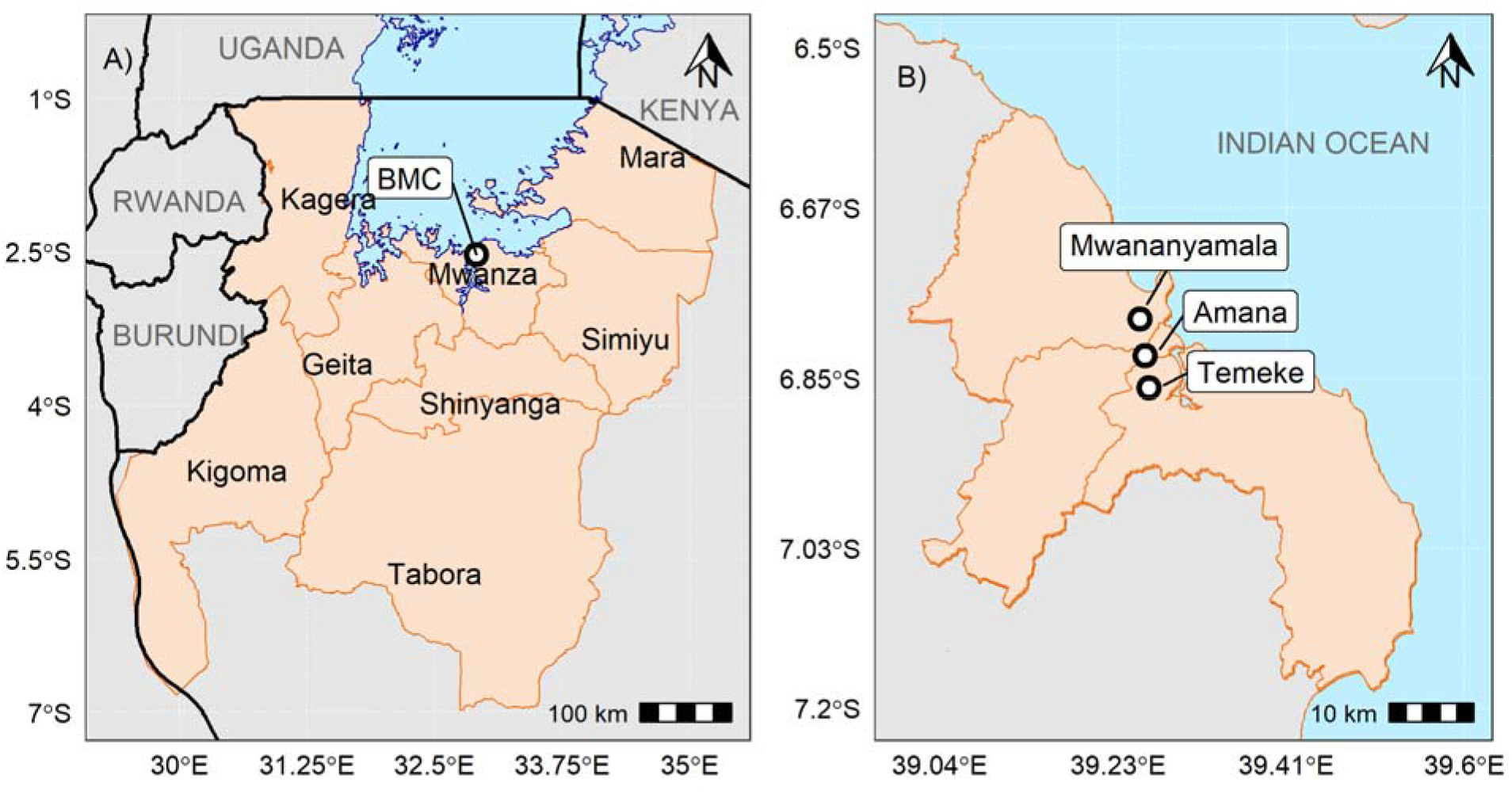
A) is a map showing the eight regions (Mara, Kagera, Mwanza, Simiyu, Geita, Shinyanga, Kigoma and Tabora) in Tanzania served by the Bugando Medical Centre in Mwanza as a zonal referral hospital Figure 2 B) is a map of Dar es Salaam showing the districts served by the three regional hospitals in Dar es Salaam - ARRH, MRRH and TRRH.

### 3.2 Study Population

Forty (40) individuals participated in this study from three groups of stakeholders. (i) Policy makers **(n= 4)**-included here were district medical officer in charge, officers involved in coordinating NCDs and/ or SCD programs at the ministerial level and members of SCD taskforce at the Ministry (ii) Implementers **(n= 21)**-HCPs involved in delivering health care services related to NBS for SCD and its comprehensive care services-pediatricians, hematologists, physicians, laboratory technologists, midwifery and other nurses (iii) Families **(n= 15)**-parents and caregivers of children who took part in the screening programs for SCD. Sub-set of this data (nurses FGDs, families IDIs/FGD) was previously analyzed to understand the influence of gender norms in the settings in relation to quality of care of a child born with SCD [28]. In this paper, we have included data from both policy makers (IDIs), implementers (FGDs and IDIs) and families (IDIs and FGD) to address a different question on sustaining implementation of NBS for SCD in Tanzania.

### 3.3 Data Collection Methods

We conducted four **(4)** Focus Group Discussions (FGDs) with nurses working in postnatal and neonatal sections of three public hospitals in Dar es Salaam-ARRH, MRRH, TRRH. Fourteen nurses (n= **14)** participated in the FGDs. Since the nurses at TRRH already had experience in two previous NBS for SCD programs, the discussions were structured to learn the following; acceptability of the screening program to the families, experience in providing health education to the families before the screening, collection and storage of the samples as well as training required. On the other hand, FGDs with nurses in ARRH and MRRH were tailored to understand their perspective on the potential implementation of NBS for SCD in their facilities.

Using semi structured interview guide, we conducted four **(n= 4)** In-depth Interviews (IDIs) with policy makers to understand contribution (past, current and future) of Ministry of Health to build sustainable NBS for SCD programs and its comprehensive care services. IDIs with the HCPs **(7)** outside the nurses aim to understand their experiences or perspectives in implementation and ways that NBS can be sustained as well as in providing comprehensive care to the SCD patients. IDIs with families **(n= 15)** were tailored to learn from their own personal experience of NBS for SCD as well as when accessing care for SCD. Figure 3 below illustrate participants enrolled in the study and data collection methods.

**Figure 2.**
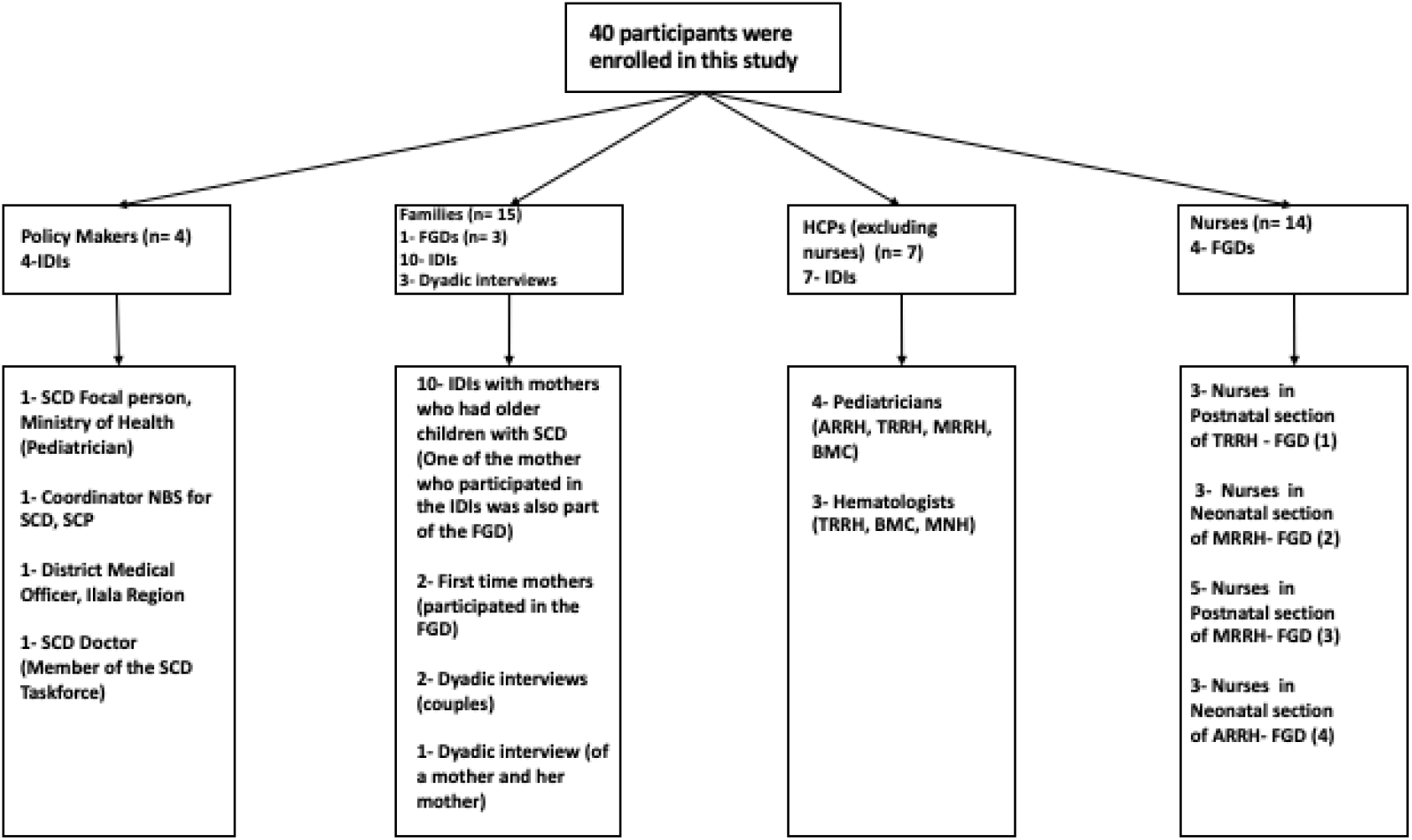
Illustrating participant enrolled in this study and data collection methods.

**Figure 3:**
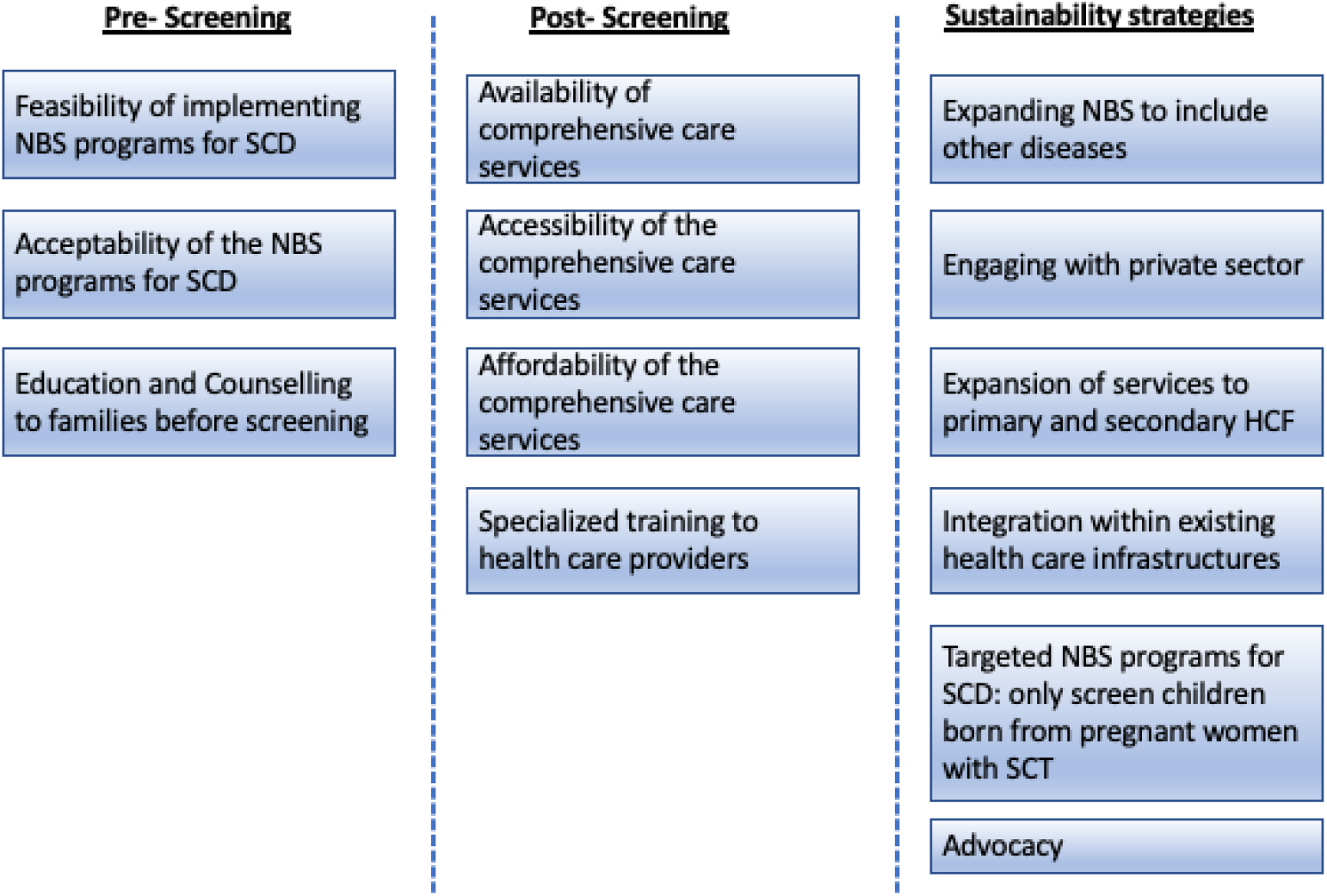
Summary of the main themes from the framework analysis grouped into; Pre-Screening, Post-Screening and Sustainability strategies.

### 3.4 Data Analysis

Thematic content analysis was used to analyze the data using framework analysis techniques to allow a more structured approach in the analysis [29, 30]. The analysis started with transcription of all forty (n= 40) interviews verbatim. Three transcripts were selected for preliminary analysis and familiarization with the interview. At this stage, one transcript was selected from each of the three groups (families, implementers and policy makers). The three transcripts were coded broadly in relation to the research questions but also allowing for any emerging issues brought about by the data. All related codes were grouped together into one theme, the themes were also grouped into distinctive categories based on whether they fall into either implementation (included here were themes related to the provision of NBS for SCD services and comprehensive care services) or sustainability (included here were themes related to proposing ways to sustain continuity of NBS comprehensive care services component of the NBS for SCD. The framework used was categorized into three main categories (i) Pre-screening: grouped here were themes that need to be considered before and during occurrence of the screening (ii) Post-screening-grouped here were themes related to the implementation of comprehensive care services for SCD (iii) Sustainability approaches-grouped here were themes related to how NBS for SCD and care can be sustained in LMICs. The themes and sub-themes were then charted and indexed into the framework analysis in relation to the three main categories. After charting and indexing all the three transcripts, the framework was shared for inputs. The feedback was incorporated into improving the framework, which was then used to map the codes for the remaining transcripts. Codes that fall outside the three main categories were not included in the analysis of the data used for this manuscript. Figure 3 below is summarizing the main themes from the analysis grouped into the three categories: Pre-Screening, Post-Screening and Sustainability strategies.

## 4.0 Results

### 4.1 Pre-screening

In this section, we highlight three main thematic topic (i) Feasibility of implementing NBS for SCD programs (ii) Acceptability of the programs in the settings (iii) Education and Counselling to families

#### 4.1.1 Feasibility of conducting NBS for SCD

Both implementers and policy makers were positive on the possibility of establishing NBS for SCD programs in regional referral hospitals (RRH) where specialized care for SCD already existed. The strategy was to start the programs in well-resourced RRH and then moving to district hospitals (DH). Challenges addressed by the clinicians as well as policy makers were on laboratory diagnostics, specialized training to the health care providers and the need for high level advocacy to ensure government support in the implementation.

> *“We have also talked about policy makers and how we can take them on board to ensure newborn screening for sickle cell disease is prioritized. If we succeed in doing that then all the activities will be budgeted by the Ministry of Health [NBS for SCD], having its own budget it will be easy for the implementers. Also, the hospital will need to have qualified personnel who are knowledgeable about the program to provide education to the families. The same with Laboratory diagnostics and other interventions. Since hematologists are very few in Tanzania, we will have to encourage our doctors to pursue those trainings and our nurses to fully implement the programs” 1_IDI_Implementers_Paediatrician_MRRH_06.08.18*

#### 4.1.2 Acceptability of NBS for SCD

Participants in all the three categories acknowledge the benefits of NBS for SCD. Nurses who were working in the programs explained that very few mothers refuse to screen their newborn babies for SCD. This observation was also consistent during the interviews with families, mothers recall that during the screening most mothers were keen to screen their newborn babies for SCD. The few refusals were linked to low knowledge of SCD. Parents who already have children with SCD were more interested to know the results and initiate requests to screen their newborn babies than those who did not have children with SCD.

> *“From our experiences of screening these children for sickle cell disease, we never had a situation where mothers will refuse to screen their babies for whatever reasons. Most of them are happy to get their children screened. We never had refusal based on certain culture [or tribe] not allowing newborn babies to be screened” 1-FGD_Implementers_Nurses_Postnatal Unit_TRRH_07.08.18*

During the discussion with nurses from Amana hospital, one of the nurses who had prior experience of working in screening programs, suggested low knowledge of the disease in the communities as potential reason that may lead to refusal to screen.

> *“Mothers may find it hard to accept because of the little understanding they have about the disease, but when you educate them then it is easy to accept screening their children. So, her own understanding will influence her to screen the child. We also tell the mothers the benefits of knowing early if your child is born with the disease” 2_FGD_Implementers_Nurses_Neonatal Unit_ARRH_27.08.18*
>
> Acceptability of the screening was also linked to having family history of the disease, as indicated during a dyad-interview with a mother and father of a child with SCD diagnosed through NBS.
>
> *“For me when I gave birth, the nurse came and advise us or may be requested us to screen our children for sickle cell disease, and she asked us, are you ready? and because I know this problem is there [in her family] I was also attracted to screen my child for sickle cell, and I say okay I am ready. They took the blood, and they promise to call us to give us the result back” 5_IDI_Families_both parents_30.01.2019*

#### 4.1.3 Education and Counselling to families

In hospitals with ongoing screening services, nurses acknowledge education and counselling need to be integrated in the services. Lack of adequate number of nurses in the postnatal wards limit time that nurses may use for educating families. Suggestion were made for health education to start at the antenatal clinics to ensure mothers are educated about SCD before the actual screening. Another challenge raised during the interviews was nurses themselves felt that they do not have enough knowledge on SCD to competently and comprehensively provide counselling and health education to the families.

> “*Education to nurses is needed for them to provide counselling to the families. It is difficult to counsel or educate another person if you don’t have enough knowledge of the disease” 7_IDI_Implementers_Haematologist_TRRH_14.09.18*

In the group interview with mothers who participated in the screening program, they pointed out the need to have detailed education session on SCD. Since the health education included other issues related to child-care the SCD part was felt by the mothers to be very small.

> *“However, the education provided was a small bit of everything, we are educated not only about sickle cell even how to breastfeed our babies. Therefore, we did not think of even asking questions on sickle cell-M1 (Probe: so, when do you think is the right time to provide the education) Education has to start in the antenatal clinics, because the day when we gave birth, the mother is being told so many things, you cannot remember everything” 1_FGD_Families_TRRH_01.09.18*

### 4.2 Post-screening

For NBS program to be effective it has to be complemented with comprehensive care services. Under this section, we have identified four key thematic topics related to comprehensive care services for SCD in Tanzania (i) Availability of comprehensive care services (ii) Accessibility of the comprehensive care services (iii) Affordability of the comprehensive care services (iv) Specialized Training to health care providers.

#### 4.2.1 Availability of comprehensive care services

All the RRH that participated in this study have specialized clinics for SCD at least once per week. The average number of patients consulted in the clinics’ ranges from 20 to 32 weekly. The clinics are attended by nurses, doctors specialized in providing care for sickle cell patients and/ or hematologist and pediatrician. The sickle cell clinics are well integrated services through the existing health care infrastructure. The key issue highlighted throughout the discussion was a need to establish sickle cell clinics in the primary health care facilities and availability of diagnostic tests.

> *“The most challenging aspect is diagnostic tests, but sickle cell clinics have been there in some of the regional referral hospitals, and we do have data in terms of the number of patients visiting those clinics for over the years” 1-IDI_Policy makers_ Coordinator of SCD programs, Ministry of Health_20.08.2018*

#### 4.2.1 Accessibility of comprehensive care services

Since most of the specialized care for SCD are in regional referral hospitals as illustrated in *figure 4* below. Families are required to travel fairly a long distance to access care. Lack of money for transport was linked to poor clinic attendance for some families. One of the suggestions provided by nurses who closely work with the families was to introduce services in health care facilities in primary and secondary and build capacity to provide specialized care for SCD. This solution will also redistribute the burden of care so that the RRHs provide specialized care for SCD who are not managed at primary and secondary health facilities. Families were also concerned by the level of care provided at primary and secondary health care facilities and reiterated the suggestion to provide trainings on specialized care for SCD at all levels of health care.

**Figure 4.**
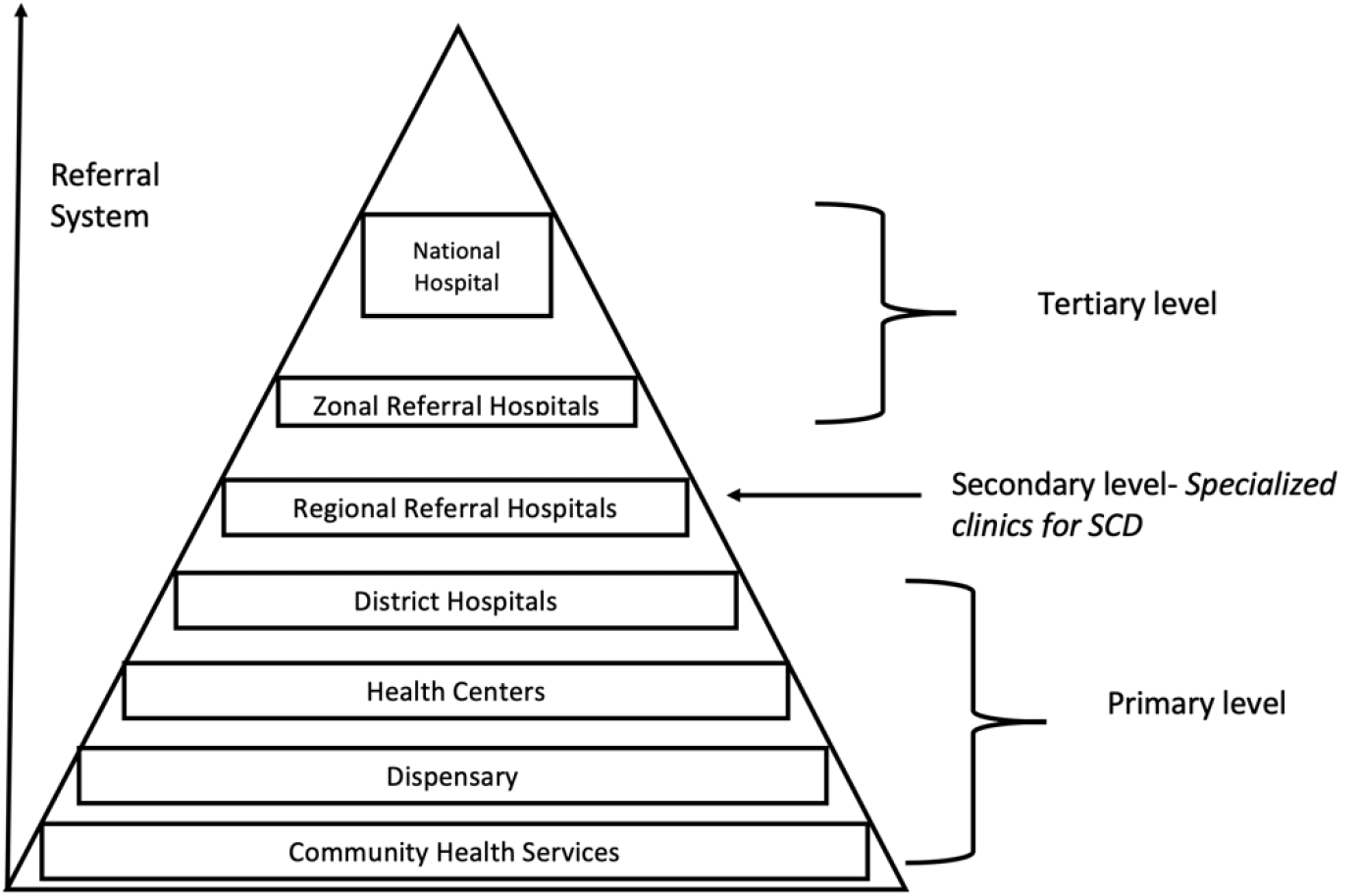
Illustration of Health Service Levels and Referral system in Tanzania.

> *“It is also important to do follow ups with patients if they are not attending clinics, but if we just wait for patients to come to the clinic [without follow ups] then there are many patients who cannot come to clinics because they don’t have money for transport. The child may be sick, and they will not be able to bring them to the hospital. Temeke is so big people are coming from different places and if all the children will have to come to Temeke, it is not going to be possible” 1-FGD_Implementers_Nurses_Postnatal Unit_TRRH_07.08.18*

#### 4.2.3 Affordability of the health care services

According to the government policy, children under five years of age and those suffering from chronic illnesses such as SCD, can access health care for free. This policy has enabled most of the families to bring their children to the clinics for care. However, implementers acknowledge the challenges in the implementation of the exemption policies. Inconsistencies on the availability of the essential medicines like Penicillin and Folic acid require parents to sometimes pay for the medicines out of pockets. The National Health Insurance Fund (NHIF) that is also considered as one of the possible solutions to subsidize health care costs for children under 18 years of age. At the time of the interviews, all the thirteen (13) families interviewed in this study their children were not insured by the NHIF.

> *“[P1] The government policy require children with chronic illness to access care for free, so for pediatric sickle cell clinic, drugs are given for free. I am not sure if Penicillin is available [they might be asked to go and buy) but folic acid is given for free. [P2] We encourage families to buy insurance for their children, most parents believe that if you have children under five then everything is free, but it is not everything, insurance will be able to pay so many things” 4_FGD_Implementers_Nurses_Neonatal Unit_MRRH_09.08.18*

During one of the family interviews, there were concerns that not all parents will be able to buy insurance package for their children, suggestion was made to treat sickle cell patients for free.

> *“With regard to the services, we just need to have the health insurance, for those who don’t have insurance they are getting so many difficulties, for example the other day when I came to the clinic, that thing for taking out blood [Cannula?], I was told to go and buy it, sometime there are no drugs you have to pay out of pocket, so those are some of the challenges. Some families don’t have money to pay for the services or buy the insurance. The only solution is for sickle cell patients to be treated for free” 11_IDI_Families_15.02.2019*

#### 4.2.4 Trainings for HCPs and Human Resources for Health Crises

Training in genetic counselling, diagnostic methods and management of SCD were considered to be important by the implementers and policy makers. Nurses acknowledged that once they receive the right training in SCD and genetic counselling, they will be better equipped to educate families.

*“Training are still needed, on our side [nurses] we need to be trained before we start educating families. If we are knowledgeable about the disease, the benefits of screening and the management options available, then it will be easy to educate the families” 3_FGD_Implementers_Nurses_Postnatal Unit_MRRH_09.08.18*

The interview with the head of pediatric unit in one of the RRH emphasized the importance of creating sustainable training plans. Instead of focusing on individuals, the training should be tailored to benefit majority of the staff in the respective units.

> *“Since this service is new then, the nurses will need refresher training for those who will be responsible for running the programs. And also we should not select one or two people to get the training, because other staff will wait until that person is around to do the work [the program become this person project] or telling the families come another day the [nurse] is not around. So if it will include postnatal unit then all the nurses need to be trained [how to screen, how to transport the sample and the diagnostic aspect] it has to be integrated with the hospital structure. If you use one or two person you will need to pay them and that will bring segregation amongst the staff” 5_IDI_Implementers_Paediatrician_TRRH_13.08.18*

### 4.3 Proposed sustainability strategies that may be applicable to low resources settings

#### 4.3.1 Expanding NBS to include other diseases

Healthcare providers and policy makers both agreed that expanding NBS services for SCD to include other conditions, could be one of cost-effective approach to leverage the limited resources. Suggested newborn disorders that can be added to include HIV screening, hearing disorders, down syndrome and other physical abnormalities such as talipes.

> *“Expanding the screening services will give us an opportunity to do so many tests under one umbrella, so the same DBS samples can be used to screen other diseases such as HIV. Now, the question is do we have the resources to do all the tests? but as a recommendation I think we should think of it [expanding] the issues of resources will come after we are ready. The primary aim is to benefit the child” IDI_DMO_Ilala_Policymaker_20.08.18*

Similar suggestion was also provided during the FGDs with nurses at TRRH

> *“This needs to be advocated to the ministry [expanding NBS programs] to involve other diseases as a way of minimizing the costs for screening instead of just screening for one disease. Although this feels like a very long-term plan” 2_FGD_Implementers_Nurses_Neonatal Unit_ARRH_27.08.18*

Bugando hospital in Mwanza did a pilot project to screen SCD through the DBS sample collected for HIV screening. However, since newborn screening for HIV is only done for children born with HIV positive mothers only fewer groups will be able to benefit.

> *“We did a pilot program to screen all newborn babies for sickle cell disease born in Bugando hospital centers but due to issues of reagents we had to stop. And we did start another program to screen only a small population of children born with exposed mothers [children who were less than 24 months] since all the DBS are coming to our hospitals from all regions, we thought this is a very good platform to screen for Sickle Cell Disease. We have completed this project three weeks ago, and I can say the prevalence is still very high” 3_IDI_Implementers_Paediatrician_BMC_15.08.18*

#### 4.3.2 Engaging with private sector

Partnering with private health facilities and diagnostic laboratories to sustain implementation of the screening programs appear to be a feasible sustainable approach. The Hematology Clinical and Research laboratory (HCRL-Public) at MUHAS partnered with the Aga Khan hospital (AKH-Private) in Dar es Salaam in processing NBS samples. The costs paid by AKH allow the laboratory to sustain itself through buying reagents and also subsidize the testing costs for samples from public health facilities.

> *“At the ministry of health, we have a section that only deals with public private partnership and also another section that deals with private health facilities and also diagnostic service department. Through those resources we can assess in what ways we can benefit from each other especially in the area of diagnosis. We are aware that private health facilities are doing a number of screening, so yes there is a lot that we can learn and leverage from each other” 2_IDI_MoHGEC_Policy Maker_01.08.18*

#### 4.3.3 Expansion of services to primary and secondary HCF

Providing essential training to HCPs working in primary health care facilities was suggested as a strategy to sustain the comprehensive care services for SCD. This will help to distribute the workload across the different levels of care and can potentially reduce the burden of care in regional referral hospitals and zonal hospitals.

> *“We are hoping for the best on the newborn screening for Sickle Cell Disease, whatever we are doing there [in the referral hospitals] can also be done in primary and secondary health care facilities. We can start piloting the programs in regional referral hospitals like Temeke, it is still possible to collect DBS in primary as well as secondary health care facilities with much higher number of births. Sample can be collected daily and processed in a centralized laboratory [Muhimbili] Nurses that have been trained here in Temeke can train other nurses in the health facilities” 5_IDI_Implementers_Paediatrician_TRRH_13.08.18*

#### 4.3.4 Integration within existing health care infrastructures

Except for BMC, NBS for SCD programs are currently implemented in public health hospitals. HCP, pharmacies and laboratories are supported by the government. The only gap that the NBS program is fully covering is related to laboratory diagnosis. In-service training was provided to nurses and clinicians working in those public hospitals. The specialized clinics for SCD are part of the pediatrics clinics for the hospitals.

> *“If we want these services [NBS for SCD] to be sustainable it has to be part of the hospital plans, it has to be one of the services offered in the hospital and not a funded program that when the donors leave then the project is not there. If it will be part of the hospital services, then it can be sustained even at a lowest scale possible. This means when the hospital is doing budgeting if there are any reagents or laboratory diagnostics then will be budgeted” 2_FGD_Implementers_Nurses_Neonatal Unit_ARRH_27.08.18*

#### 4.3.5 Targeted NBS programs for SCD: only screen children born from pregnant women with SCT

Other proposed suggestions to sustainably implement NBS for SCD in limited resource settings was to only screen those at-risk children born through parents with SCT. Health care workers believed that this approach will be more cost effective in lowering down the screening costs. Mothers who have sickle cell trait will be captured through the antenatal clinic. Screening for SCD at birth will only be done to newborns whose mothers have positive test results. This model is similar to that adapted for HIV screening at birth.

> *“Families where mothers will be identified to have sickle cell trait, then the children will have to be screened. The antenatal card can be used to capture this information [this is within our ability] On my side I will take this forward and start implementing” 1_IDI_DMO_Ilala_Policymaker_20.08.18*

#### 4.3.7 Advocacy

There are two types of advocacy which is being done. The first level of advocacy is with patients’ groups to build awareness and provide education to the public on SCD. The contribution of these campaigns is having a community which is knowledgeable about the disease and willing to allow children to be screened for the disease. The second level of advocacy is with government officials and policy makers.

> *“We are doing advocacy campaigns targeting the community as well as the decision makers [government officials, policy makers]. For example, the school campaigns program that we are currently involved to build awareness amongst secondary school children. With regard to the government and Ministry, we are trying to provide evidence on how big this problem is and what are the possible solutions. We do believe with continuous advocacy we can achieve our goals” Member of the SCD taskforce, Ministry of Health “So again, that’s a policy issue and I think that’s why for me it’s almost impossible to talk about anything in isolation because we need multiple initiatives at the same time. So, while we are doing NBS we are also supposed to be talking to the Government about what we are doing [through providing evidence], our role should be advising– for what works and what doesn’t. If it has to be sustainable at some point, we need to see the Government owning the program. So how can that be? I think it’s going to be a very slow process; the most important thing is we are going in that direction” IDI_Implementers _NBS for SCD Coordinator_Sickle Cell Program_07.022019*

## 5.0 Discussion

In this paper we have shared experience and perspectives of families, HCPs and policy makers in the implementation of NBS for SCD programs as well as provision of comprehensive care for SCD patients in Tanzania. While success in the implementation was limited, there are significant efforts that have been done locally to ensure continuity of care even at the lowest scale possible. Although we have used experience of NBS for SCD in Tanzania to model the potential opportunities and challenges to sustain the programs, the experiences outlined in this paper can be learned and adapted in other screening programs implemented in limited resource contexts. Opting for the structured analysis assisted us to outline our results and discussion based on the key themes emerging from this study.

### Pre-screening

Assessing acceptability of health intervention programs implemented in a new setting is one of the key steps to measure overall success of a program [31, 32]. Evidence already exist in the literature on the different kinds of social-cultural and religious beliefs around SCD in Africa that can be linked with poor uptake of the screening programs [33–37]. However, in this case acceptability was not considered a challenge by both families and HCPs involved in this study. Our study did not find evidence to show that the few refusals to screen were related to any cultural beliefs in any of the tribes or religions. The results were consistent to surveys that were designed to investigate acceptability of NBS for SCD in different zones of Nigeria [17, 38]. The sub-set of this data that was analyzed to understand how gender norms influences quality of care of the child with SCD, reveals disproportionate burden of care to the mothers [28]. The findings drive the discussion beyond not just acceptability but also understanding social norms that may interfere with the child’s quality of care after screening [39].

In terms of feasibility, both pilot NBS for SCD program in Tanzania that were implemented in Dar es Salaam and Mwanza provided baseline data on the burden and feasibility of implementing screening programs [16, 40]. Challenges highlighted by the HCPs were in diagnostic equipment and specialized training in laboratory and SCD management. Several other pilot NBS for SCD programs highlighted similar challenges in other parts of Africa [12, 14, 17, 18, 20, 41]. Point of care testing (POCT) has been recommended as the most cost effective solution for diagnosis of SCD in places with high burden of the disease and limited resources [22, 42]. Data from pilot studies on POCT implemented in Africa demonstrated both affordability and reliability of the tests [24, 43]. Collectively, this data provides strong evidence on potential of POCT to be standard practice for SCD diagnosis with the aim of expanding screening coverage.

### Post screening

Both the pilot screening program in Dar es Salaam and Mwanza were implemented in hospitals with existing clinics offering specialized care for SCD [16, 40]. This strategic approach ensured all children diagnosed are immediately enrolled into available comprehensive care services. Since the specialized care is currently available in RRHs, suggestions were made to expand to primary and secondary health care facilities. This will spare families from travelling a long distance for care and at the same time will reduce the burden of care from the RHH. Our study on gender and care for a child with SCD has shown how mothers are disproportionately affected with care of the child [28]. By bringing care closer to families this will facilitate clinic attendance and help reduce the burden of care to women. Another advantage of this approach is to start building capacity of hospitals at the primary and secondary levels to provide specialized care for SCD. This approach will require a need assessment; first, to identify potential facilities located in areas where prevalence of SCD is high. Secondly, leveraging into existing clinics that can also be used as sickle cell clinics at least once by week. Commitment from the regional and district hospital authorities is key to ensure sustainability of the care to patients. Capacity building include providing relevant training to nurses and clinicians already working in the hospitals, improving laboratory infrastructures and development of management guidelines and relevant policies for SCD.

In facilitating accessibility and affordability of the health care provided families were encouraged by HCPs to enroll children into health insurance programmes. The National Health Insurance Fund (NHIF) in its mission to expand its coverage to reach out to the most disadvantageous categories has developed a subsidized package for children under 18 years of age costing around 22 dollars per year. This package has enabled more children from low income families to access health care services up to tertiary level. During the discussions with nurses at TRRH they did indicate that only few children coming to the hospital have health insurance. In most families, men are working and will be required to support the health care costs since mothers are looking after the sick children. If the father is unsupportive most likely will not be willing to incur the health care costs [28]. This observation is concerning and may require understanding of family’s attitude and perspectives on budgeting for health care related costs.

### Sustainability strategies

Establishing screening programs in existing programs and health care infrastructure was recommended by the implementers and policy makers as one of the sustainability strategies. The pilot NBS for SCD that was implemented through immunization program in Nigeria provided evidence on the feasibility and sustainability of the model in limited resources [44]. Leveraging the limited resources through working with other developed programs in the settings has also been recommended through other experiences [4, 23, 45]. Integration of the screening into developed HIV programs was also suggested in this study as a strategy to leverage the limited resources. Others have also made a similar approach to use HIV programs in Africa as a platform to append other screening programs [15]. Although this suggestion seems plausible a cost effective analysis is needed to inform sustainability of this model in limited resources [2, 46].

One of the suggestions provided by the policy makers was to explore Public Private Partnerships (PPP). Ministry of Health in Tanzania has a dedicated section dealing with PPP that can facilitate opportunities to partner with private sector to strengthen screening program. The challenges in most of the public health facilities are in laboratory diagnostics. Partnering with private laboratory diagnostic facilities to develop a shared laboratory diagnostic center can address the challenge [42]. The partnership between HCRL at MUHAS as a public health facility and Aga Khan hospital in Dar es Salaam as a private facility is a good case to model how PPP can support NBS for SCD in limited resources [47–49].

The proposal for NBS for SCD to target only families with sickle trait in low resources settings has also been suggested elsewhere [50]. Focusing on screening mothers during antenatal clinics and identify those with traits will reduce the costs to screen all children for SCD even those who are born from parents without the trait.

The contribution of advocacy is considerably important in driving health services allocation at both global and local level [15, 47]. Advocacy campaigns aimed to educate families and public on SCD and related public health interventions have played a big role in Tanzania to create awareness on the disease. Patients groups like SCYF, SCPCT, TASIWA and TANSCDA continue to develop different platforms to communicate messages on SCD to the public and the government. Part of the recognized contribution of advocacy in Tanzania include inclusion of Sickle Cell Disease in the National Strategic Planning of Non-Communicable Diseases as well as inclusion of Hydroxyurea as essential medicines to SCD Patients.

## 6.0 Conclusion

Developing and sustaining newborn screening programs in countries in Africa is challenging due to competing public health priorities and limited health care budgets. NBS for SCD programs in Tanzania has demonstrated both the opportunities and areas that need addressing s in the implementation and sustainability of the services in low resource settings. A key area of strengthening is full integration of the services in health care. However, efforts at the local level to sustain the programs are encouraging and can be used as model in other programs implemented in low resources settings.

## Data Availability

Data sharing not applicable as no datasets generated and/or analysed for this study. All data relevant to the study are included in the article.

## 7.0 Ethical Consideration

The study was approved by the Muhimbili University of Health and Allied Sciences Research Ethics Committee (Ref. No. 2017-10-20/ AEC/ Vol. XII/85). Consent was sought from all participants prior to the conduct of the study.

8.0 Acknowledgements

Authors are grateful for the time of the families, health care workers and policy makers who participated in this study

## 9.0 Funding

DB research support was through student small grant at MUHAS. Commonwealth Scholarship Commission funded the DB to spend 1 year at Ethox centre where analysis and initial writing of this work started.

## 10.0 Competing Interest

Authors declare no conflict of interests

## 11.0 Authors contribution

DB designed the study, interacted with participants, performed interviews, assisted in transcriptions and translations, analysis and drafting of the manuscript. JM and MP supervised DB and reviewed the drafts of the manuscripts. SM provided technical inputs in the analysis plan and reviewing the manuscript. SN, CM, KM assisted in revising the manuscript. All authors reviewed and approved the final manuscript.

## 12.0 Patient Consent for publication

Not required

## 13.0 Patient and public involvement

Patients and/or the public were not involved in the design, or conduct, or reporting or dissemination plans of this research.

## Abbreviations

AfriSickleNet: Africa Sickle Cell Research Network
ARRH: Amana Regional Referral Hospital
BMC: Bugando Medical Centre
DHH: District Health Hospital
FGDs: Focus Group Discussions
GAVI: Global Alliance for Vaccine and Immunization
HCRL: Hematology Clinical Research Laboratory
HIV: Human Immunodeficiency Virus
HCP: Health Care Providers
HCF: Health Care Facilities
IDIs: In- Depth Interviews
LMICs: Low Middle Income Countries
MNH: Muhimbili National Hospital
MRRH: Mwananyamala Regional Referral Hospital
NBS: Newborn Screening
POCT: Point of Care Testing
RRH: Regional Referral Hospitals
SCD: Sickle Cell Disease
SCP: Sickle Cell Program
SCPCT: Sickle Cell Patients Community of Tanzania
SPARCO: Sickle Pan African Research Consortium
SYCF: Sickle Cell Youth Foundation
TANSCDA: Tanzania Sickle Cell Disease Alliance
TASIWA: Tanzania Sickle Cell Warriors
TRRH: Temeke Regional Referral Hospital

## Notes

### Competing Interest Statement

The authors have declared no competing interest.

### Author Declarations

The study was approved by the Muhimbili University of Health and Allied Sciences Research Ethics Committee (Ref. No. 2017- 10-20/ AEC/ Vol. XII/85). Consent was sought from all participants prior to the conduct of the study.

